# Community’s misconception about COVID-19 and its associated factors: Evidence from a cross-sectional study in Bangladesh

**DOI:** 10.1101/2021.04.12.21254829

**Authors:** Md. Bakebillah, Md. Arif Billah, Befikadu L. Wubishet, Md. Nuruzzaman Khan

**Affiliations:** Department of Folklore, Faculty of Social Science, Jatiya Kabi Kazi Nazrul Islam University, Trishal-2224, Mymensingh, Bangladesh; Department of Psychology and Counselling, Faculty of Business, Economic and Social Development, Universiti Malaysia Terengganu, 21030 Kuala Nerus, Terengganu, Malaysia; Postdoctoral Research Fellow, Health Services Research Centre, Faculty of Medicine, The University of Queensland, Brisbane Qld 4072 Australia; Department of Population Science, Faculty of Social Science, Jatiya Kabi Kazi Najrul Islam University, Trishal-2224, Mymensingh, Bangladesh

**Keywords:** Misconception, social and mass media, Rural area, COVID-19, Bangladesh

## Abstract

**Introduction:** Misconception about COVID-19 has been spread out broadly that the World Health Organization declared it as a major challenge in the fight against the disease. This study aimed to assess common misconceptions about COVID-19 among the rural people of Bangladesh and its associated socio-demographic and media related factors.

**Methods:** Data were collected from 210 respondents selected from three unions of Satkhira District, Bangladesh.The dependent variable was misconception about COVID-19 (Yes, No) which was generated based on the respondents’ responses to six questions on common misconceptions of COVID-19. Explanatory variables were respondents’ socio-demographic characteristics, mass media and social media use habits. Descriptive statistics were used to describe the characteristics of the respondents. Univariate and multivariate logistic regression models were used to determine the factors associated with misconception about COVID-19.

**Results:** Misconceptions about COVID-19 were found among more than half of the total respondents. More than 50% of the respondents reported they consider COVID-19 as a punishment from God. Besides, most of the respondents reported that they do not think COVID-19 is dangerous (59%), and COVID-19 is a disease (19%). Around 7% of the total respondents reported they consider this virus as a part of a virus war (7.2%). Bivariate analysis found that socio-demographic characteristics of the respondents and factors related to social and mass media were significantly associated with misconception. However, when all the factors included together in the multivariate model, the likelihood of misconception was lower among secondary (AOR, 0.33, 95% CI, 0.13-0.84) and tertiary (AOR, 0.29, 95% CI, 0.09-0.92) educated respondents compared to respondents with primary education.

**Conclusion:** This study obtained a very higher percentage of the rural people of Bangladesh having one or more misconceptions related to COVID-19. This could be a potential challenge in the fight against the pandemic. Effective use of mass and social media to communicate evidence based information on COVID-19 as well as to educate the public about COVID-19 is important.

## Background

Since December 2019, the world has been faced with the coronavirus outbreak which the World Health Organization later declared as a pandemic. All countries in the world are now affected by the pandemic and recorded high numbers of cases and deaths. Specially older people and individuals with pre-existing comorbidities such as diabetes and hypertension have been disproportionately affected by the pandemic. [1]. Similar to previous pandemics such as HIV/AIDS and Ebola, COVID-19 has been surrounded by a number of misconceptions from the very beginning [10,11] The existence of widely held misconceptions related to COVID-19, particularly in the remote regions of developing countries including Bangladesh, has also been an important challenge compromising COVID-19 preventive efforts in communities [2,3]. For instance, many religious groups believe that this virus mainly affects non-religious communities or atheists while others believe educated and richer people are more prone to get the infection [3,4]. People in low-income countries have also been observed to believe that they have better immuity than people in developed countries which prevent them from getting COVID-19 [5,6]. There are other groups that believe coronavirus is a revenge of nature and punishment from God [7–9]. These misconceptions are dangerous because they deter disease preventive behaviours and also people with such misconceptions are less likely to report any cases and seek for health care [3,7,9]. The scarcity of evidence based information on both the virus and the disease as well as rapid emergence of these misinformation from numerous sources including social media are major reasons why the public dwelled in an “ocean” of misconceptions for so long [12]. Unfortunately misconeptions and misinformation are disseminated much quicker the correct information. The WHO defines this situation as ‘*infodemic’*, rapid and far-reaching spread of both accurate and inaccurate information, which can potentially affect mental health and health related behavior of societies [12–14].

Bangladesh is one of the countries worst hit by the COVID-19 pandemic. More than a half million people has been infected among which around 9,000 people were died as per the official estimates of the World Health Organization for Bangladesh [15]. Given the low testing capacity of facilities and lack of accurate report of causes of deaths, these numbers could be reflecting underestimation of the real cases and deaths in the country, which is reflected in the reports of several private organizations and surveys [16,17]. Moreover, many Bangladeshi people have been observed to hold misconceptions about this virus, which makes them hide even if they have been infected or even death of a family member [16]. Another cause of possible underreporting of the true infection and death rates of this outbreak is social pressure. People mostly experience social isolation if they have been infected and therefore they prefer to hide their diagnosis. Importantly, these sorts of behaviors are common among people in rural and remote areas than their counterparts residing in urban areas [17–19].

There is a lack of study in Bangladesh on the issue of misconception about COVID-19. Only a few studies have been conducted but with several limitations including inappropriate list of misconceptions and inadequate sample [20,21]. Other focus areas of previous studies have been knowledge and awareness about COVID-19 among the general population [22], health professionals [23,24], parents [25] and students [26] and vulnerable and susceptible populations [27–29]. Moreover, very few studies have been conducted to explore misconceptions about COVID-19 that were disseminated through social and mass media [5,30,31]. Evidence from the rural area is scarce till now. Therefore, this study aimed to explore the COVID-19 related misconception among the rural people of Bangladesh.

## Methodology

### Data source and sample

Primary data was collected during the period April 15 to May 09, 2020. Data was collected from three unions (smallest administrative classification of Bangladesh), Ratanpur, Kusulia and Moutala Union) of Satkhira district, Bangladesh. Multistage sampling was employed to select the unions and villages included in the study. At the first stage of the selection, Satkhira district was selected randomly from list of 64 districts in Bangladesh. These three unions have 68 villages [32]. We did a further random sampling to select 9 villages from these, three villages from each union. There were 113 households of the selected 9 villages of which 105 households were included in this survey. A fixed number of two members were included from each household with the inclusion criteria, (i) the youngest person aged more than 18 and (ii) the oldest person. The criteria were choosen to present family’s most (over age 18) youngest and oldest person. This produced responses from 210 respondents which were our analysis sample for the study (Figure 1).

**Figure 1.**
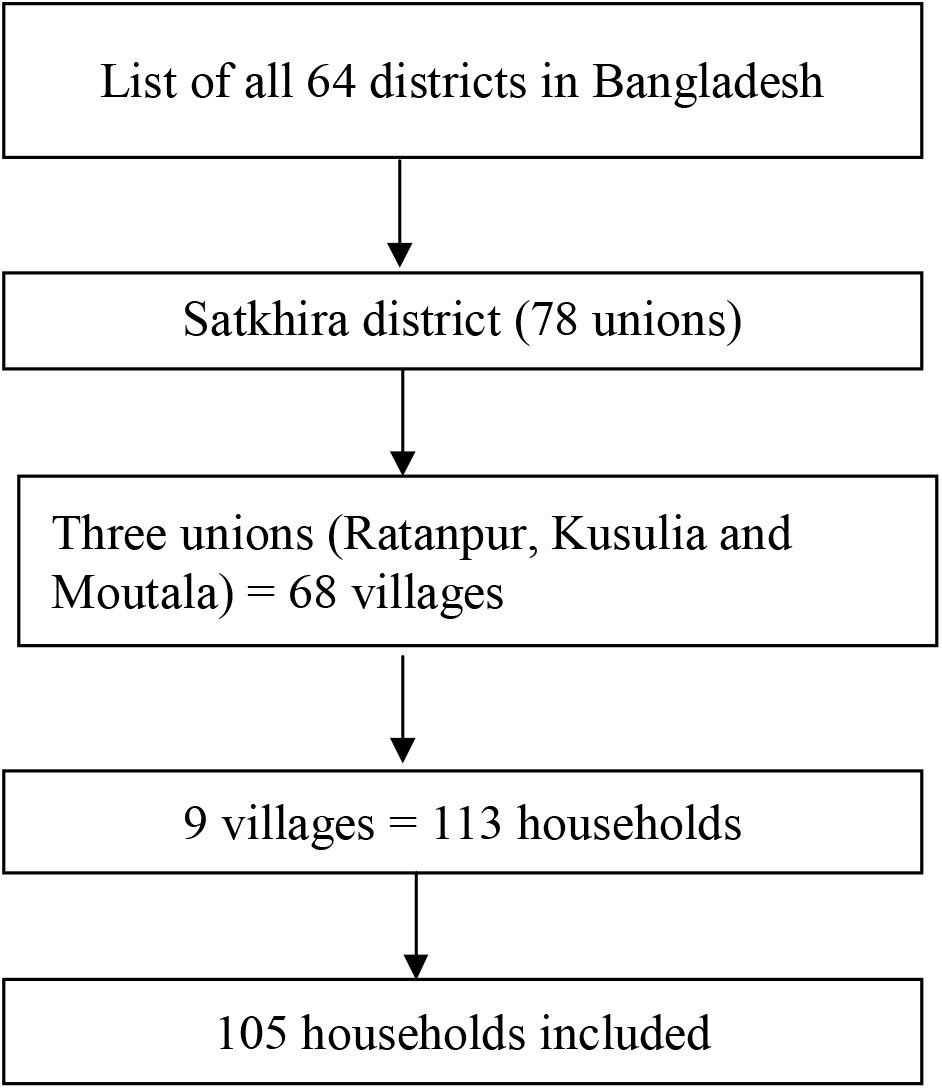
Sampling procedure for the study

### Data collection

The questionnaire was first prepared in English and translated into Bengali, official language of Bangladesh. It was then back translated into English, to make sure the original meaning was retained. The questionnaire was developed based in the evidence of relevant research in Bangladesh and other developing countries [3,5,21,22,33,34]. Two rounds of pre-testing were conducted, and inaccuracies and misunderstood questions were revised before the final data collection. Verbal consent was taken from each respondent before collecting the data.

#### *Dependent* variable

The dependent variable was misconception about COVID-19, that was created based on the respondent’s response to six forms of misconception which were reported in Bangladesh at the time of the survey. These were COVID-19 is: (i) a punishment from god, (ii) revenge of nature, (iii) a virus war, (iv) not a dangerous virus, (v) not a disease and (vi) not a pandemic. Response to each statement was reported dichotomously, “Yes” *or* “No”. We used these responses to generate a misconception variable with two categories, respondents had no misconception (0, if given correct response to all statements) and respondents had misconception (1, if given incorrect response to any of these statements).

### Explanatory variables

The explanatory variables included were respondents’ socio-demographic characteristics and social and mass-media related practices. Socio-demographic characteristics included respondents’ gender, age, education and monthly income. Social and mass media related variables included respondents use of social media, daily frequency of social media use, reading newspapers, frequency of reading newspaper, watching television, frequency of watching television, listening to radio and frequency of listening to radio.

### Analysis

Descriptive statistics were used to describe the characteristics of the respondents. Chi-square test was used to identify the variables significantly associated with misconception as well as their distribution. Bivariate and multivariate logistic regression models were used to identify the factors associated with misconception. The variables found significant in the chi-square analysis at 10% level of significance were included in the multivariate logistic regression model. Results were reported as Odd Ratio (OR) with 95% confidence interval (CI). The statistical software STATA version 14.3 was used for all statistical analyses.

## Results

### Characteristics of the study population

The characteristics of the respondents are presented in Table 1. Around two-thirds (66.19%) of the respondents were male. Most of the respondents (53.81%) were middle-aged (30-60 years) and completed their secondary education (43.81%). The mean household income where the respondents resided in was 15,645 (SE, 758.123) Bangladeshi Taka (BDT). Most of the respondents lived in a small family (57.61%) and 67.14% had children in the family. About 87% of them reported use of mass media, of which 41% read newspapers, 82.4% watch television and 6.2% listen to the radio at least once in a week. Around 60% of the total respondents reported use of social media, more than a quarter of which using social media for more than two hours per day.

**Table 1.**
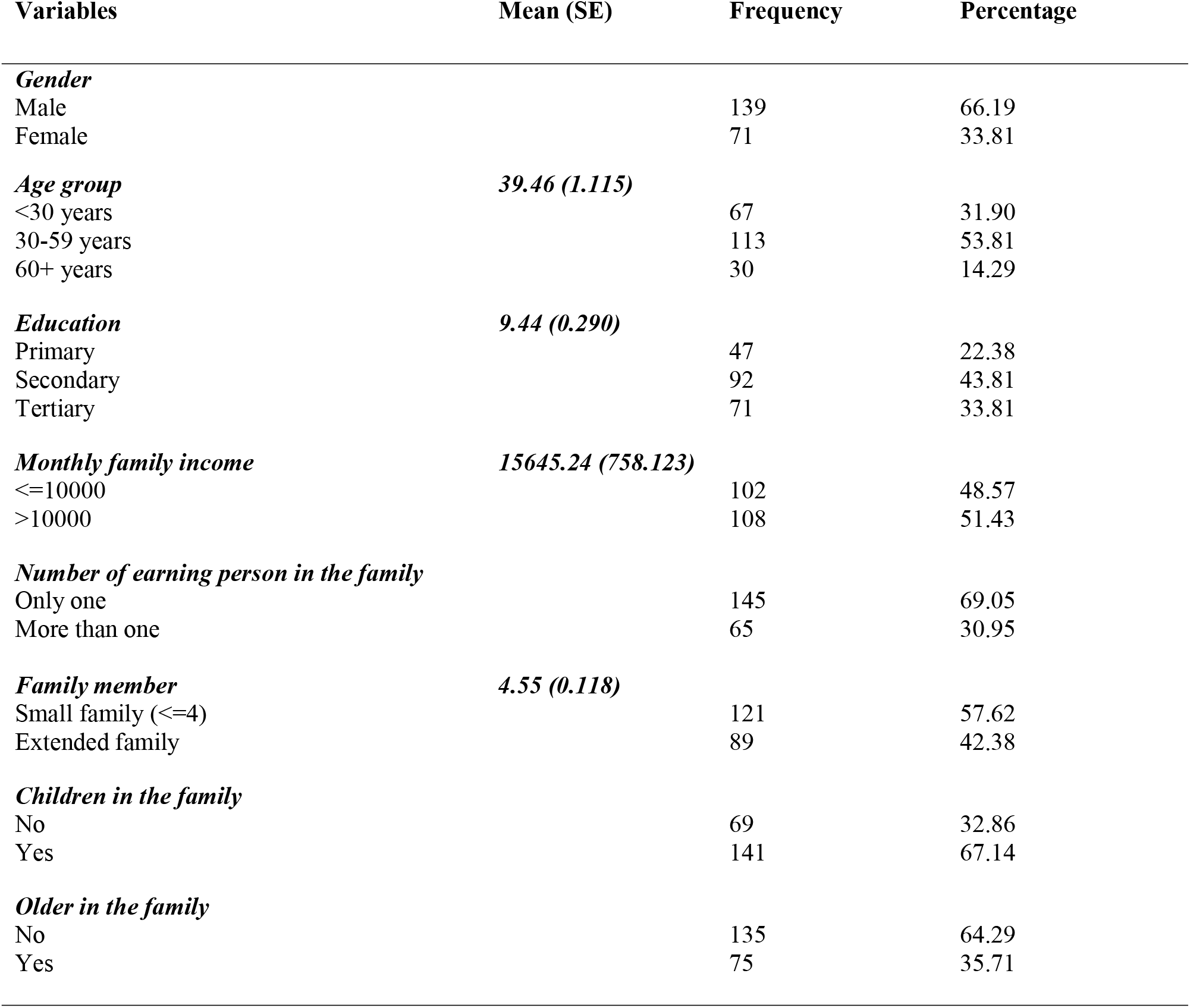
Household characteristics of the study population

| **Variables** | **Mean (SE)** | **Frequency** | **Percentage** |
| --- | --- | --- | --- |
| ***Gender*** |  |  |  |
| Male |  | 139 | 66.19 |
| Female |  | 71 | 33.81 |
| ***Age group*** | **39.46 (1.115)** |  |  |
| <30 years |  | 67 | 31.90 |
| 30-59 years |  | 113 | 53.81 |
| 60+ years |  | 30 | 14.29 |
| ***Education*** | **9.44 (0.290)** |  |  |
| Primary |  | 47 | 22.38 |
| Secondary |  | 92 | 43.81 |
| Tertiary |  | 71 | 33.81 |
| ***Monthly family income*** | **15645.24 (758.123)** |  |  |
| <=10000 |  | 102 | 48.57 |
| >10000 |  | 108 | 51.43 |
| ***Number of earning person in the family*** |  |  |  |
| Only one |  | 145 | 69.05 |
| More than one |  | 65 | 30.95 |
| ***Family member*** | **4.55 (0.118)** |  |  |
| Small family (<=4) |  | 121 | 57.62 |
| Extended family |  | 89 | 42.38 |
| ***Children in the family*** |  |  |  |
| No |  | 69 | 32.86 |
| Yes |  | 141 | 67.14 |
| ***Older in the family*** |  |  |  |
| No |  | 135 | 64.29 |
| Yes |  | 75 | 35.71 |

Common misconception among the study population are presented in Table 2. More than half (52.86%) of the respondents reported one or more misconceptions. The most popular misconception in the study population was not considering the COVID-19 virus as dangerous (59%), followed by the belief that COVID-19 is a punishment from God (47%). There were also respondents who do not believe that the virus may cause a disease (19%) and some believe that COVID-19 is a virus war (7.6%).

**Table 2:** Common COVID-19 related misconceptions among rural people in Bangladesh, 2020

| Attributes | Frequency | Percentage |
| --- | --- | --- |
| COVID-19 is a punishment from God |  |  |
| Have no misconception | 111 | 52.9 |
| Have misconception | 99 | 47.1 |
| COVID-19 is the revenge of nature |  |  |
| Have no misconception | 205 | 97.6 |
| Have misconception | 5 | 2.4 |
| COVID-19 is a virus war |  |  |
| Have no misconception | 194 | 92.4 |
| Have misconception | 16 | 7.6 |
| COVID-19 is a dangerous virus |  |  |
| Have no misconception | 87 | 41.1 |
| Have misconception | 123 | 58.6 |
| COVID-19 is a disease |  |  |
| Have no misconception | 171 | 81.4 |
| Have misconception | 39 | 18.6 |
| COVID-19 is a pandemic |  |  |
| Have no misconception | 209 | 99.5 |
| Have misconception | 1 | 0.5 |

The distribution of respondents with or without misconception across socio-demographic and media related factors is presented in Table 3. Misconception was reported higer among men (74.65%) than their women counterpart (25.25%). Lower level of misconception was found among the respondents who had secondary (47.47%) or tertiary (42.42%) level of education. About 70.27% respondents who did not read newspaper had high level of misconception whereas 87.88% respondents who watched television had low level of misconceptions about the COVID-19 disease. Besides, those who use social media (72.73%) had a higher level of misconception regarding COVID-19 compared to the 52.25% misconnection among sample who reported they use social media. We found gender, monthly income, television use, treatment facilities, remedy measures were significantly associated with COVID-19 related misconception (p<0.05) in the bivariate analysis.

**Table 3.** Socio-demographic and media related variables among respondents with and without the COVID-19 related misconception

| Variable | COVID-19 related misconception |  | Chi-square/Fisher exact test (df), p-value |
| --- | --- | --- | --- |
|  | No | Yes |  |
| ***Gender*** |  |  | ***6.1286 (1), 0.013*** |
| Male | 74 (74.65) | 65 (58.56) |  |
| Female | 25 (25.25) | 46 (41.44) |  |
| ***Education*** |  |  | ***17.3052 (2), 0.000*** |
| Primary | 10 (1.11) | 37 (33.33) |  |
| Secondary | 47 (47.47) | 45 (40.54) |  |
| Tertiary | 42 (42.42) | 29 (26.13) |  |
| ***Monthly family income*** |  |  | ***5.0016 (1), 0.025*** |
| <=10000 | 40 (40.40) | 62 (55.86) |  |
| >10000 | 59 (59.60) | 49 (44.14) |  |
| ***Read Newspaper*** |  |  | ***11.2862 (1), 0.001*** |
| No | 47 (47.47) | 78 (70.27) |  |
| Yes | 52 (52.53) | 33 (29.73) |  |
| ***Watch Television*** |  |  | ***3.9004 (1), 0.048*** |
| No | 12 (12.12) | 25 (22.25) |  |
| Yes | 87 (87.88) | 86 (77.48) |  |
| ***Listen Radio*** |  |  | ***2.7132 (1), 0.100*** |
| No | 90 (90.91) | 107 (96.40) |  |
| Yes | 9 (9.09) | 4 (3.60) |  |
| ***Frequency of reading newspaper*** |  |  | ***10.6437 (2), 0.005*** |
| Not read | 46 (46.46) | 76 (68.47) |  |
| Read but not daily | 13 (13.13) | 7 (6.31) |  |
| Read daily | 40 (40.40) | 28 (25.23) |  |
| ***Frequency of watching Television*** |  |  | ***4.9216 (2), 0.085*** |
| Not watch | 12 (12.12) | 26 (23.42) |  |
| Watch but not daily | 5 (5.05) | 7 (6.31) |  |
| Watch daily | 82 (82.83) | 78 (70.27) |  |
| ***Frequency of listening radio*** |  |  | ***3.0914 (2), 0.213*** |
| Not listen | 90 (90.91) | 107 (96.40) |  |
| Listen but not daily | 5 (5.05) | 3 (2.70) |  |
| Listen daily | 4 (4.04) | 1 (0.90) |  |
| ***Use social media*** |  |  | ***13.5524 (1), 0.000*** |
| No | 27 (27.27) | 58 (52.25) |  |
| Yes | 72 (72.73) | 53 (47.75) |  |
| ***Frequency of using social media/ day*** |  |  | ***14.4197 (2), 0.001*** |
| Don't use | 27 (27.27) | 58 (52.25) |  |
| <=2 hrs | 36 (36.36) | 31 (27.93) |  |
| >2 hrs | 36 (36.36) | 22 (19.82) |  |
| ***Treatment facility*** |  |  | ***7.5754 (2), 0.023*** |
| No treatment | 45 (45.45) | 53 (47.75) |  |
| Non-specialized treatment | 16 (16.16) | 32 (28.83) |  |
| Specialized treatment | 38 (38.38) | 26 (23.42) |  |
| ***Remedy*** |  |  | ***6.8192 (2), 0.033*** |
| Prayer | 7 (7.07) | 14 (12.61) |  |
| Awareness | 41 (41.41) | 28 (25.23) |  |
| Both | 51 (51.52) | 69 (62.16) |  |
| ***Role of social media*** |  |  | ***16.5407 (2), 0.000*** |
| Negative | 8 (8.08) | 6 (5.41) |  |
| Positive | 78 (78.79) | 63 (56.76) |  |
| NA | 13 (13.13) | 42 (37.84) |  |

Results from the unadjusted and adjusted multivariate logistic regression analyses are presented in Table 4. Unadjusted logistic analysis found higher likelihood of misconception among women (OR, 2.1, 95% CI, 1.16 - 3.77) compared with men. Respondents who had reported their monthly income as more than 10000 BDT were 46% (OR, 0.54, 95% CI, 0.31 - 0.93) less likely to have misconception compared with those who reported their monthly income to be less than 10000 BDT (p<0.01). Compared with the primary educated respondents, those who attained secondary and tertiary level of education were 74% (05% CI, 0.12-0.58) and 81% (95%CI, 0.08-0.43) less likely to have misconception, respectively. Besides, the use of mass media (0.26, 0.10-67), and social media ≤2 hrs per day (0.40, 0.21-0.78) and >2 hrs per day (0.28, 0.14-0.57) were found to be associated with 74%, and 60% and 72% lower level of misconception, respectively.

**Table 4.**
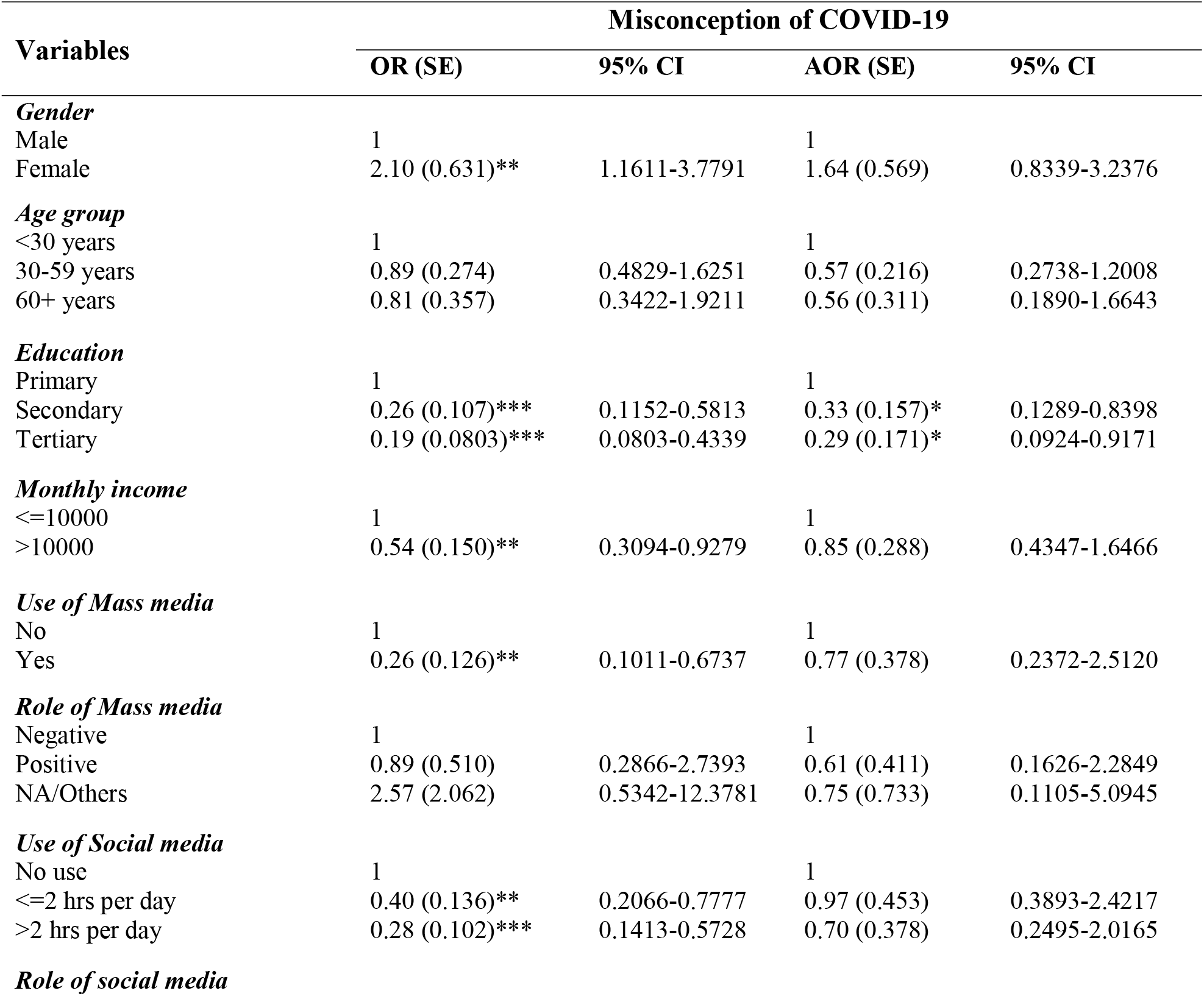
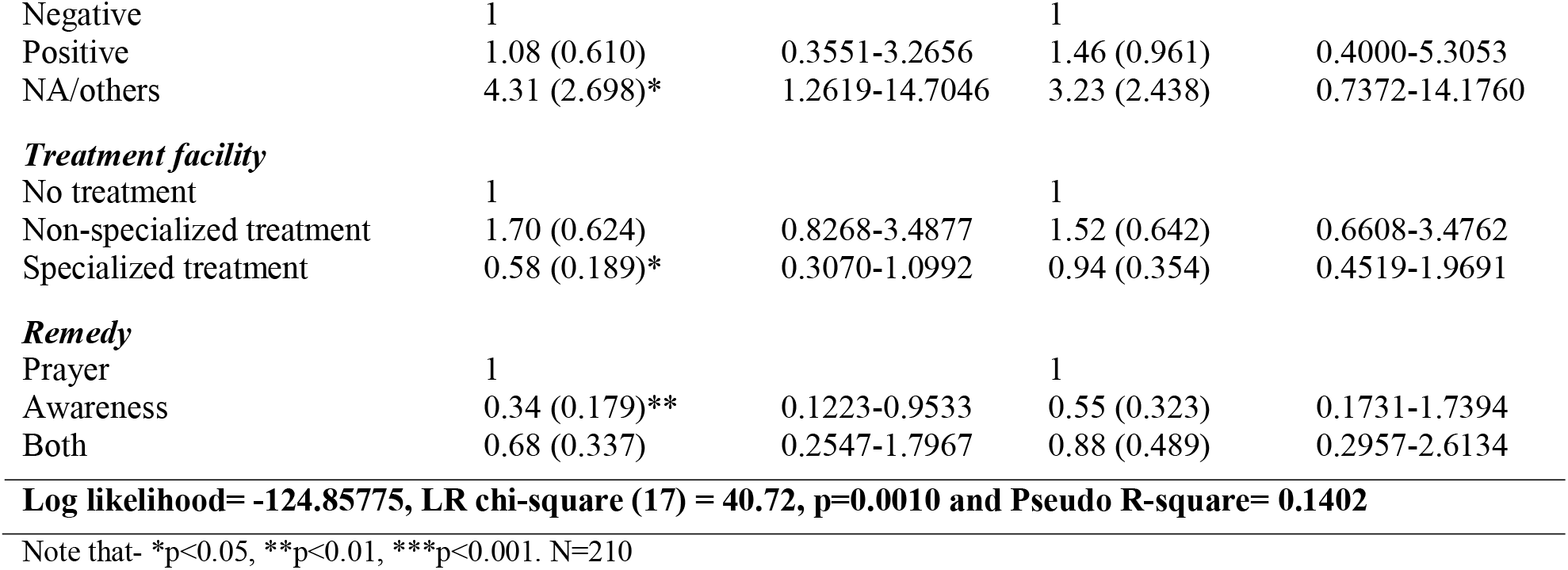
Factors associated with the misconception about COVID-19 in Bangladesh

| Variables | Misconception of COVID-19 |  |  |  |
| --- | --- | --- | --- | --- |
|  | OR (SE) | 95% CI | AOR (SE) | 95% CI |
| **Gender** |  |  |  |  |
| Male | 1 |  | 1 |  |
| Female | 2.10 (0.631)** | 1.1611-3.7791 | 1.64 (0.569) | 0.8339-3.2376 |
| **Age group** |  |  |  |  |
| <30 years | 1 |  | 1 |  |
| 30-59 years | 0.89 (0.274) | 0.4829-1.6251 | 0.57 (0.216) | 0.2738-1.2008 |
| 60+ years | 0.81 (0.357) | 0.3422-1.9211 | 0.56 (0.311) | 0.1890-1.6643 |
| **Education** |  |  |  |  |
| Primary | 1 |  | 1 |  |
| Secondary | 0.26 (0.107)*** | 0.1152-0.5813 | 0.33 (0.157)* | 0.1289-0.8398 |
| Tertiary | 0.19 (0.0803)*** | 0.0803-0.4339 | 0.29 (0.171)* | 0.0924-0.9171 |
| **Monthly income** |  |  |  |  |
| $\leq 10000$ | 1 | | 1 | |
| $> 10000$ | 0.54 (0.150)** | 0.3094-0.9279 | 0.85 (0.288) | 0.4347-1.6466 |
| **Use of Mass media** |  |  |  |  |
| No | 1 |  | 1 |  |
| Yes | 0.26 (0.126)** | 0.1011-0.6737 | 0.77 (0.378) | 0.2372-2.5120 |
| **Role of Mass media** |  |  |  |  |
| Negative | 1 |  | 1 |  |
| Positive | 0.89 (0.510) | 0.2866-2.7393 | 0.61 (0.411) | 0.1626-2.2849 |
| NA/Others | 2.57 (2.062) | 0.5342-12.3781 | 0.75 (0.733) | 0.1105-5.0945 |
| **Use of Social media** |  |  |  |  |
| No use | 1 |  | 1 |  |
| $\leq 2$ hrs per day | 0.40 (0.136)** | 0.2066-0.7777 | 0.97 (0.453) | 0.3893-2.4217 |
| $> 2$ hrs per day | 0.28 (0.102)*** | 0.1413-0.5728 | 0.70 (0.378) | 0.2495-2.0165 |
| **Role of social media** |  |  |  |  |
| Negative | 1 |  | 1 |  |
| Positive | 1.08 (0.610) | 0.3551-3.2656 | 1.46 (0.961) | 0.4000-5.3053 |
| NA/others | 4.31 (2.698)* | 1.2619-14.7046 | 3.23 (2.438) | 0.7372-14.1760 |
| ***Treatment facility*** |  |  |  |  |
| No treatment | 1 |  | 1 |  |
| Non-specialized treatment | 1.70 (0.624) | 0.8268-3.4877 | 1.52 (0.642) | 0.6608-3.4762 |
| Specialized treatment | 0.58 (0.189)* | 0.3070-1.0992 | 0.94 (0.354) | 0.4519-1.9691 |
| ***Remedy*** |  |  |  |  |
| Prayer | 1 |  | 1 |  |
| Awareness | 0.34 (0.179)** | 0.1223-0.9533 | 0.55 (0.323) | 0.1731-1.7394 |
| Both | 0.68 (0.337) | 0.2547-1.7967 | 0.88 (0.489) | 0.2957-2.6134 |
| **Log likelihood= -124.85775, LR chi-square (17) = 40.72, p=0.0010 and Pseudo R-square= 0.1402** |  |  |  |  |

Note that- \*p<0.05, \*\*p<0.01, \*\*\*p<0.001. N=210

Adjusting the confounders, only education of the respondents had a significant association with misconception about COVID-19. Respondents having secondary and tertiary education were found 0.33 (95% CI, 0.13-84) and 0.29 (95% CI 0.09-0.92) less likely to report misconception about COVID-19 respectively than those having primary education. Importantly, the factors related to social and mass media were not as significant predictors of COVID-19 related misconceptions once respondent’s characteristics were adjusted.

## Discussion

The study identified common misconceptions related to COVID-19 in Bangladesh and their association with factors related to sociodemographic, mass and social media use of respondents. We found a high level of misconception about COVID-19 among our study sample which vary significantly across respondents’ socio-demographic and media related factors. We also reported that the use of social and mass media and increase education were significantly related to the lower level of misconception about COVID-19 in rural Bangladesh. These findings are robust and could have an important policy implication to reduce the misconception over this pandemic in Bangladesh and other LMICs.

This study reported a very high level of misconception with around 52% of the total respondents. The commonest forms of misconception were this pandemic is a punishment of nature and the coronavirus (the virus that is known to cause COVID-19) is not a dangerous virus. These types of misconception were promoted by religious leaders and covered in several social and mass media [4,8]. Moreover, though it is established that COVID-19 is spill over infection and not a human made (laboratory generated) output [35–37], however, another common misconception is that this virus is human created biological weapons for international politics [38]. Similar findings were reported across the world, including America [9,39], Saudi Arabia [31] Nigeria [33,40], Nepal [7], Ghana [41], Uganda [42] and Pakistan [43].

Socio-demographic characteristics of the respondents were associated with COVID-19 related misconceptions. For instance, COVID-19 related misconceptions are more common among people living in rural areas as reported in a previous study in Bangladesh [22]. People living in rural areas are usually have lower education and have limited access to the healthcare facilities and mass medias that could contribute to more widespread presence of such misconceptions. This is particularly true for the rural women of Bangladesh, as they are even more family-centric and most have lower education which is usually associated with higher infectious disease burden and related misconceptions. For instance, a significant proportion of the rural women believe children’s respiratory disease (Acute respiratory diseases) was the influence of evils or supernatural beings, and therefore can only be treated through the spiritual healers [44,45]. Though these sorts of misconception are different from the misconceptions related to the COVID-19, however, their existence work as an easy root to spread out the COVID-19 related misconception.

Education plays a significant role to prevent infectious disease, like COVID-19, through better aware people creation about disease, education on causes of infection and ways of prevention. Educated people are usually have better access to the mass and social media which are the comments roots of spreading COVID-19 related misconceptions. However, educated people have the capacity to differentiate between the right and wrong. This enables them to recived the correct information from the mass and social media and avoid incorrect information which contribute to the lowering misconceptions [43,46]. Another root of lower misconception about the COVID-19 among the educated people is access to the advanced medical treatment facilities (43).

The Government of Bangladesh is now using mass media to reduce misconceptions about the ongoing COVID-19 pandemic and disseminate correct information. The WHO recommends the same strategy to fight against this pandemic. For this, the newspapers, television channels have been broadcasting the regular updates about the spread of the disease, new findings, national and international strategies. However, access to mass media, except the radio, is significantly lower in rural area of Bangladesh compared to the urban area. The effectiveness of radio to disseminate such information has been questioned for a variety of reasons. For instance, radio often disseminate the one way of information, which is mostly difficult to understand for the rural and uneducated people. Moreover, such one way of information dissemination can have the little influence to justify and reduce misconception that the rural people get from other sources including social media where unauthentic information spread out rapidly. Additionally, it mostly fails to answer the questions that people have had regarding this disease, thorough this is an important way out to reduce misconception. Ensuring proper access to the mass media and dissemination of correct knowledge could therefore have a significant role to reduce misconception about COVID-19 pandemic.

This study has number of strength and limiatations. As far we know, this is the first study of its kind in Bangladesh that assess the misconception regarding the COVID-19 pandemic and associated factors. Moreover, this study consider a wide range of factors and identify their association with the COVID-19 related misconception. This makes this study findings is more reliable. However, data collected was cross-sectional in nature, thereofore this study findings is correlational only, not causal. Moreover, this study data was collected from only one district and three unions, therefore they findings may not be geralizable for the whole population of Bangladesh though the district and unions included were selected randomly.

## Conclusion

We found that many people living in rural parts of Bangladesh have one or more misconceptions about COVID-19. Respondents’ socio-demographic characteristics and status of using mass and social media were identified as significant predictors of misconception related to COVID-19. Proper education and dissemination of correct information in mass and social media are important to reduce the misconceptions and succeed in the fight against this pandemic.

## Data Availability

The data will be made available following reasonable request to the corresponding author.

## Acknowledgement

We acknowledged to the respondents who had participated in this study voluntarily. We also give thanks to Mst. Sumaiya Sultana (Post Graduate student), Sk Mustafizur Rahman (physiotherapist), Prokash Patra (Science teacher at a high school), Ajgor Ali (Post graduate student), and Jayanta Kumar Ghosh (Village doctor) for being associated during the data collection at the field level. We also acknowledged the Department of Folklore, Faculty of Social Science, Jatiya Kabi Nazrul Islam University, Trishal-2224, Mymensingh, Bangladesh where this study is conducted.

## Abbreviations

COVID-19: Coronavirus Disease 2019
AOR: Adjusted Odds Ratio
OR: Odds ratio
SE: Standard Error
CI: Confidence Interval
HIV/AIDS: Human Immuno-deficiency Virus/ Acquired Immunodeficiency Syndrome
WHO: World Health Organization

